# Atrophy of the Nucleus Basalis of Meynert predicts the progression of gait variability in Parkinson’s disease

**DOI:** 10.1101/2020.05.12.20099523

**Authors:** Kevin B. Wilkins, Jordan E. Parker, Helen M. Bronte-Stewart

## Abstract

Parkinson’s disease (PD) is a systemic brain disorder where the cortical cholinergic network begins to degenerate early in the disease process. Readily accessible, quantitative, and specific behavioral markers of the cortical cholinergic network are lacking. Although degeneration of the dopaminergic network may be responsible for deficits in cardinal motor signs, the control of gait is a complex process and control of higher-order aspects of gait, such as gait variability, may be influenced by cognitive processes attributed to cholinergic networks. We investigated whether swing time variability, a metric of gait variability that is independent from gait speed, was a quantitative behavioral marker of cortical cholinergic network integrity in PD. Twenty-two individuals with PD and twenty-nine age-matched controls performed a validated stepping-in-place (SIP) task to assess swing time variability off all therapy. The PD cohort also underwent structural MRI scans to measure gray matter volume of the Nucleus Basalis of Meynert (NBM), the key node in the cortical cholinergic network. Swing time variability was also measured ON subthalamic nucleus (STN) deep brain stimulation (DBS) in PD individuals. A subset of eleven individuals with PD completed the SIP task again off all therapy after three years of continuous DBS. Clinical motor assessments were performed for each condition. Swing time variability was significantly greater (i.e., worse) in PD compared to controls and greater swing time variability was related to greater atrophy of the NBM. STN DBS significantly improved cardinal motor signs but did not improve swing time variability. Swing time variability worsened in PD, off therapy, after three years of continuous STN DBS, and NBM atrophy predicted the degree of increase. In contrast, cardinal motor signs did not progress. These results demonstrate that swing time variability is a reliable marker of cortical cholinergic health, and support a framework in which higher-order aspects of gait control in PD are reliant on the cortical cholinergic system, in contrast to other motor aspects of PD that rely on the dopaminergic network.

## Introduction

Neuronal degeneration and Lewy body formation manifest in the basal forebrain early on in the progression of Parkinson’s disease (PD), specifically in the Nucleus Basalis of Meynert (NBM) (Nakano and Hirano, 1984; Rogers et al., 1985). Close to 90% of cell makeup of the NBM is cholinergic and it serves as the primary source of cholinergic innervation to the cortex (Mesulam et al., 1983; Mesulam, 2013). Atrophy of the NBM leads to major reductions in cortical acetylcholine (ACh) levels (Johnston et al., 1979; Mufson et al., 1986), and has been associated with a wide range of cognitive deficits in PD, as well as with the worsening of cognition over time (Barrett et al., 2019; Gang et al., 2020; Pereira et al., 2020; Ray et al., 2018; Schulz et al., 2018). Medications to enhance cortical ACh are usually only implemented in later stages of PD when the cortical cholinergic tone may be so low that these are largely ineffective. This missed therapeutic strategy may be partly due to the lack of behavioral biomarkers of cortical cholinergic integrity.

Gait impairment and freezing of gait (FOG) affect nearly 75% of individuals with advanced PD (Panisset, 2004). Gait variability is associated with FOG and both have been linked to deficits in attention switching, visuospatial processing, and aspects of executive function (Amboni et al., 2013, 2008; Nantel et al., 2012; Yao et al., 2017), which have all been shown to be influenced by the cortical cholinergic system (Ballinger et al., 2016; Bohnen et al., 2006; Gratton et al., 2017; Klinkenberg et al., 2011). This suggests that the NBM-cortical cholinergic system may play a role in higher-order aspects of gait control, such as gait variability in PD, but direct evidence for this is lacking. There is some evidence that the cholinergic system may be related to gait speed in PD (Bohnen et al., 2013; Rochester et al., 2012). However, gait speed is also linked to dopaminergic networks since it and other lower order aspects of gait control such as step amplitude are consistently improved by dopaminergic medical therapy and subthalamic nucleus (STN) deep brain stimulation (DBS) (Blin et al., 1991; Roper et al., 2016; Yttri and Dudman, 2018). In contrast higher-order aspects of gait control, such as gait variability, tend to be more resistant to dopaminergic treatment, which may explain the oft observed progressive decline in gait variability in PD since current therapies do not address deficits related to the cholinergic system (Blin et al., 1991; Smulders et al., 2016).

The goal of this study was to determine whether higher order aspects of gait, such as gait variability, may be useful markers of the health of the cortical cholinergic network in PD. We questioned whether the structural integrity of the NBM was related to the degree and longitudinal progression of gait variability in PD. We specifically chose swing time variability as a measure of gait variability since it is a metric that is independent from gait speed, unlike stride and step time variability (Beauchet et al., 2009; Callisaya et al., 2010; Frenkel-Toledo et al., 2005). We hypothesized that atrophy of the NBM would be associated with greater impairment of swing time variability and that this relationship would not be seen in a similar sized brain structure, the amygdala, which is not part of the cortical cholinergic network. As NBM atrophy occurs early in PD, we hypothesized that swing time variability would be greater in moderate stage PD compared to age-matched controls. Additionally, we hypothesized that STN DBS would improve cardinal motor signs reliant on the dopaminergic system, but would not improve swing time variability, due to its reliance on the cortical cholinergic system. If these hypotheses were supported, we anticipated that swing time variability would continue to decline despite successful, continuous STN DBS, and that the degree of decline would be related to the amount of atrophy of the NBM. Meanwhile, cardinal motor signs would be slower in their decline due to the efficacy of STN DBS in targeting the dopaminergic system.

## Materials and Methods

### Human Subjects

Twenty-three individuals (16 males, 7 females; age: 59.2 ± 10.1; Table 1) with clinically established Parkinson’s disease, as part of a larger trial using a first-generation investigative sensing neurostimulator (Activa™ PC+S, Medtronic PLC, FDA Investigational Device Exemption (IDE) approved), and twenty-nine age-matched controls (16 males, 13 females; age: 62.5 ± 8.0) participated in this study (Supplementary Table 1). Individuals with PD were part of a study cohort, who underwent bilateral implantation of DBS leads (model 3389, Medtronic., Inc) in the sensorimotor region of the STN. Description of the preoperative selection criteria and surgical technique are provided in the supplementary materials. Post-operative CT scans were acquired as part of the standard clinical protocol (Brontë-Stewart et al., 2010). All experimental testing was done in the off-medication state for individuals with PD: long- and short-acting medications were withdrawn over 24 to 48 and 12 hours, respectively, and DBS therapy was turned off for at least 15 minutes before testing began for all non-DBS conditions (Trager et al., 2016). No participants were taking cholinergic medications. All participants gave written informed consent to participate in the study, which was approved by the Stanford University Institutional Review Board.

**Table 1.** **Patient Demographics**

| Patient | Age<br>(Years) | Sex | Time Since<br>Diagnosis<br>(Years) | Pre-Op<br>UPDRS III<br>(off meds) | Pre-Op<br>UPDRS III<br>(on meds) | UPDRS III<br>(ON STN<br>DBS, off<br>meds) |
| --- | --- | --- | --- | --- | --- | --- |
| 1* | 72.9 | M | 9.9 | 54 | 41 | 24 |
| 2** | 52.8 | M | 4.9 | 24 | 8 | 5 |
| 3** | 62.5 | M | 3.0 | 35 | 26 | 11 |
| 4** | 65.7 | M | 5.2 | 29 | 16 | 12 |
| 5** | 58.1 | M | 3.4 | 39 | 23 | 19 |
| 6** | 42.2 | M | 4.5 | 58 | 12 | 9 |
| 7* | 69.0 | M | 10.6 | 41 | 28 | 14 |
| 8** | 53.5 | M | 7.6 | 52 | 29 | 17 |
| 9 | 73.1 | F | 8.8 | 37 | 22 | 18 |
| 10** | 72.2 | M | 8.9 | 47 | 23 | 29 |
| 11** | 58.6 | F | 10.0 | 33 | 6 | 18 |
| 12* | 54.5 | M | 11.0 | 56 | 20 | 20 |
| 13 | 34.3 | M | 2.1 | 59 | 22 | 35 |
| 14* | 57.3 | M | 6.1 | 62 | 23 | 19 |
| 15** | 67.6 | F | 6.1 | 44 | 25 | 2 |
| 16* | 50.6 | F | 9.2 | 50 | 26 | 14 |
| 17* | 61.9 | M | 14.3 | 42 | 24 | 8 |
| 18** | 52.0 | F | 12.5 | 34 | 6 | 6 |
| 19 | 66.1 | M | 7.6 | 51 | 10 | 16 |
| 20** | 56.8 | F | 11.6 | 38 | 18 | 8 |
| 21 | 75.7 | F | 8.5 | 44 | 19 | 23 |
| 22* | 55.0 | M | 10.4 | 11 | 7 | 2 |
| 23* | 50.1 | M | 10.2 | 38 | 31 | 17 |
| Average | 59.2 |  | 8.1 | 42.5 | 20.2 | 15.0 |
| ± | ± | | ± | ± | ± | ± |
| SD | 10.1 |  | 3.2 | 11.9 | 8.7 | 8.1 |

Notes: \*indicates that the participant performed the SIP ON DBS. \*\*indicates that the participant performed the SIP ON DBS and completed the three-year follow-up visit.

### Experimental Protocols

#### Clinical assessment of motor and gait impairment

Clinical motor impairment was assessed using part III of the Movement Disorders Society Unified Parkinson’s Disease Rating Scale (MDS-UPDRS III) (Goetz et al., 2007). The total score was also broken down into four bilateral sub-scores: axial, tremor, rigidity, and bradykinesia. Quantitative measures of gait were captured using the stepping-in-place (SIP) task, which is a validated metric of gait impairment and freezing (Nantel et al., 2011) (Figure 1A). In the SIP task, participants completed 90 seconds of alternating, self-paced stepping on dual force plates.

**Figure 1.**
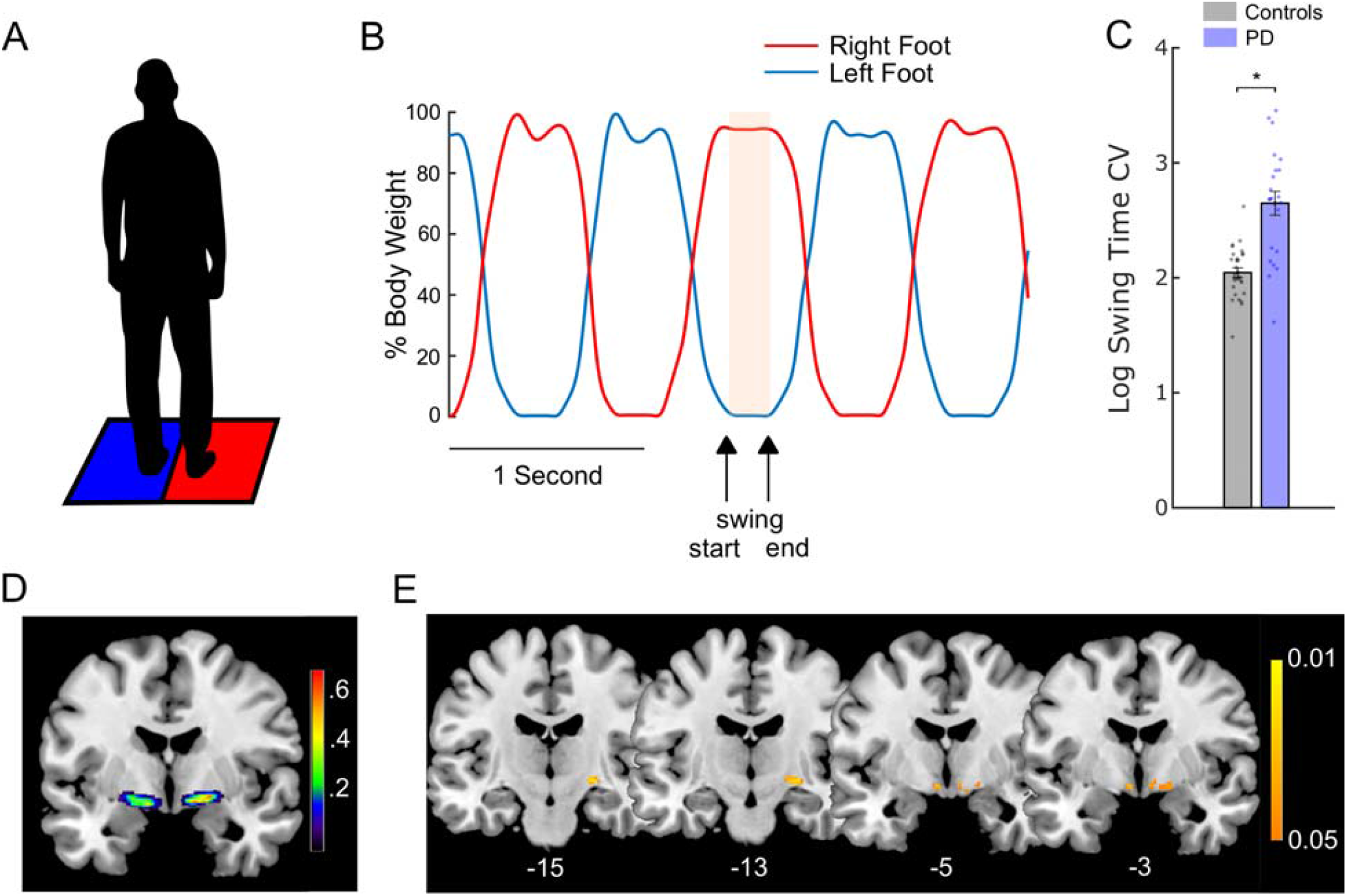
NBM atrophy was associated with greater swing time variability. (A) Depiction of the stepping-in-place (SIP) task in which the patient alternately steps on two force plates. (B) A representative trace from the SIP task showing the stepping cycles. The red line represents the stepping trace of the right foot and the blue line represents that of the left foot. The highlighted portion represents the period of time when the left foot was in the swing phase: from when the foot left the force plate to the point when it returned (arrows). The swing time coefficient of variation (standard deviation/mean) was calculated as a measure of gait variability. (C) Comparison of swing time variability between age-matched controls and the PD cohort during the SIP task. Individuals with PD showed significantly greater (i.e., worse) swing time variability. (D) Probabilistic mask of the Nucleus Basalis of Meynert (NBM) in MNI space as determined previously by Zaborszky et al., 2008. The color bar represents the probability that the voxel was identified as NBM in that cohort. (E) Voxels showing a significant negative correlation between NBM gray matter volume and swing time variability on the SIP task at Initial Programming (IP). Individuals with lower gray matter volume in the NBM show greater (i.e., worse) swing time variability. The color bar represents p-values (FWE-corrected). MNI coordinates of each slice are shown at the bottom. Error bars represent SEM. * indicates significant differences.

Age-matched controls completed only the SIP task at their visit. All PD participants completed the SIP task and were evaluated using the UPDRS III at the Initial Programming (IP) visit, prior to activation of the DBS system. A subset of nineteen individuals with PD completed the UPDRS III and the SIP task ON and OFF STN DBS at a time point between IP and the three-year visit. Lastly, a subset of eleven individuals with PD completed the UPDRS III and SIP task off all therapy after three years of continuous DBS. The breakdown of which PD individuals were included in each follow-up is denoted in Table 1.

#### Kinetic data acquisition and analysis

During the SIP task, ground reaction forces were captured at either 100 or 1000 Hz, using the Smart Equitest or Bertec systems, respectively (Neurocom Inc., Clackamas, OR, USA; Bertec Corporation, Columbus, OH, USA). Swing time was defined as the duration between when one foot left the force plate and when the same foot contacted the force plate again, Fig. 1B. Swing time variability was defined as the mean swing time coefficient of variation (CV = standard deviation/mean) and was averaged between legs. A larger swing time CV is indicative of more variable gait and has been shown to be independent of gait speed and shows high reliability (Arcolin et al., 2019; Frenkel-Toledo et al., 2005). Data were log transformed to conform to normality. A previously validated computerized algorithm was used to identify freezing episodes (Nantel et al., 2011) and swing time variability was calculated during non-freezing stepping.

#### MRI Acquisition and Analysis

Pre-operative MRI scans were acquired in the PD cohort as part of the standard clinical protocol previously described (Brontë-Stewart et al., 2010). T1-structural scans were performed at Stanford Hospital and Clinics on a 3T Discovery MR750 scanner with a 32-channel head coil. The scans had an isotropic voxel resolution of 1×1×1 mm^3^ (TR=8.24ms, TE=3.24ms, FOV= 240×240mm^2^).

#### MRI Preprocessing

Voxel-based morphometry was applied to T1 images using SPM12 software (Statistical Parametric Mapping 12; Wellcome Trust Centre for Neuroimaging, UCL, London, UK). Images were automatically segmented into gray matter, white matter, and cerebrospinal fluid with 1.5 mm^3^ resolution. The extracted gray and white matter were nonlinearly registered to Montreal Neurological Institute (MNI) space using the Diffeomorphic Anatomical Registration Through Exponentiated Lie Algebra algorithm (DARTEL). The gray matter was then warped using the individual flow fields resulting from the DARTEL registration, and voxel values were modulated for volumetric changed introduced by the high dimensional normalization to ensure that the total amount of gray matter volume was preserved. Images were then smoothed to account for anatomical variability across individuals to facilitate statistical comparison. The NBM was identified and masked using probabilistic anatomical maps available in the Anatomy Toolbox. These maps were created from histological post-mortem analysis of 10 brains (Zaborszky et al., 2008). The amygdala was used as a control region of interest from the same toolbox due to its similar size in comparison to the NBM.

#### Localization of DBS Leads

Locations of DBS leads were determined by the Lead-DBS toolbox (Horn et al., 2019) based on preoperative T1 and T2 MRIs and postoperative CT scans. Postoperative CT scans and preoperative T2 scans were co-registered to preoperative T1 scans, which were then normalized into MNI space using SPM12 and Advanced Normalization Tools (Avants et al., 2008). DBS electrode localizations were corrected for brain-shift in the postoperative CT scan (Horn and Kühn, 2015). DBS electrodes were then localized in template space using the PaCER algorithm (Husch et al., 2018) and projected onto the DISTAL Atlas to visualize overlap with the STN (Ewert et al., 2018).

#### Statistical Analysis

A Two-sample t-test was used to compare swing time variability on the SIP task in controls compared to the PD cohort at the IP visit. Voxel-wise statistics for associations between swing time variability in the SIP task in the PD cohort and gray matter volume within the NBM were computed using a general linear model in SPM12. Total intracranial volume (i.e., total white matter, gray matter, and CSF) was calculated for each individual as a global normalization to deal with brains of different sizes. A multiple regression model was used with threshold-free cluster enhancement (doi: 10.5281/zenodo.2641381) to assess any association between gray matter volume of the NBM and swing time variability in the SIP task, considering age and sex as covariates (Smith and Nichols, 2009). Significance was set at *p* < 0.05 (family-wise error corrected; FWE) with a cluster threshold of 10 voxels. The same test was run for the amygdala as a control region of interest. Paired t-tests were used to compare the effect of DBS (ON vs. OFF) on swing time variability on the SIP task, as well as change in UPDRS scores. Paired t-tests were then used to compare changes in swing time variability and UPDRS from IP to 3 years later. Significance was set at *p* < 0.05 for the t-tests. Finally, a second general linear model was computed for the percent change in swing time variability from IP to 3 years later and gray matter volume within the NBM. Due to a smaller subset for the longitudinal analysis, significance was set at p < 0.005 uncorrected with a cluster threshold of 10 voxels. The same test was run for the amygdala as a control region of interest.

## Results

### NBM Atrophy was associated with greater swing time variability

Of the full cohort of twenty-three individuals, one was excluded due to movement artifacts in their T1 scan. For the remaining twenty-two individuals, we compared swing time variability in the SIP task at the Initial Programming (IP) visit, Figure 1A and B, with age-matched controls. The PD cohort showed significantly greater (i.e., worse) swing time variability compared to controls, Fig. 1C (t(49) = 5.96, *p* = 2.69e-7).

We then compared swing time variability in the PD cohort with NBM gray matter volume, Fig. 1D, using a multiple regression model. The IP visit occurred 63.9 ± 31.9 days after the MRI scan. Multiple clusters were found bilaterally within the NBM where lower gray matter volume was significantly associated with greater (i.e., worse) swing time variability, Fig. 1E (*p* < 0.05; FWE corrected). Meanwhile, no clusters showed a positive correlation within the NBM between gray matter volume and swing time variability, nor was there any association between atrophy of the amygdala and increased swing time variability. Information on each cluster is shown in Table 2.

**Table 2.** **Correlations between NBM Gray Matter Volume and Swing Time Variability at Initial Programming**

| MNI Coordinates | Cluster Size (Voxels) | TFCE Value | p Value* |
| --- | --- | --- | --- |
| x = 24, y = -15, z = -6 | 40 voxels | 207 | $p = 0.014$ |
| x = -5, y = -5, z = -9 | 11 voxels | 169 | $p = 0.023$ |
| x = 6, y = -3, z = -9 | 36 voxels | 150 | $p = 0.029$ |

Notes: \*Family-wise error (FWE) corrected

### DBS improved cardinal motor signs but not swing time variability

Nineteen individuals were assessed with the UPDRS III and completed the SIP task OFF and ON STN DBS within the same visit at a timepoint between IP and 3 years later. Figure 2A demonstrates that the STN DBS leads were located within the STN for all individuals.

**Figure 2.**
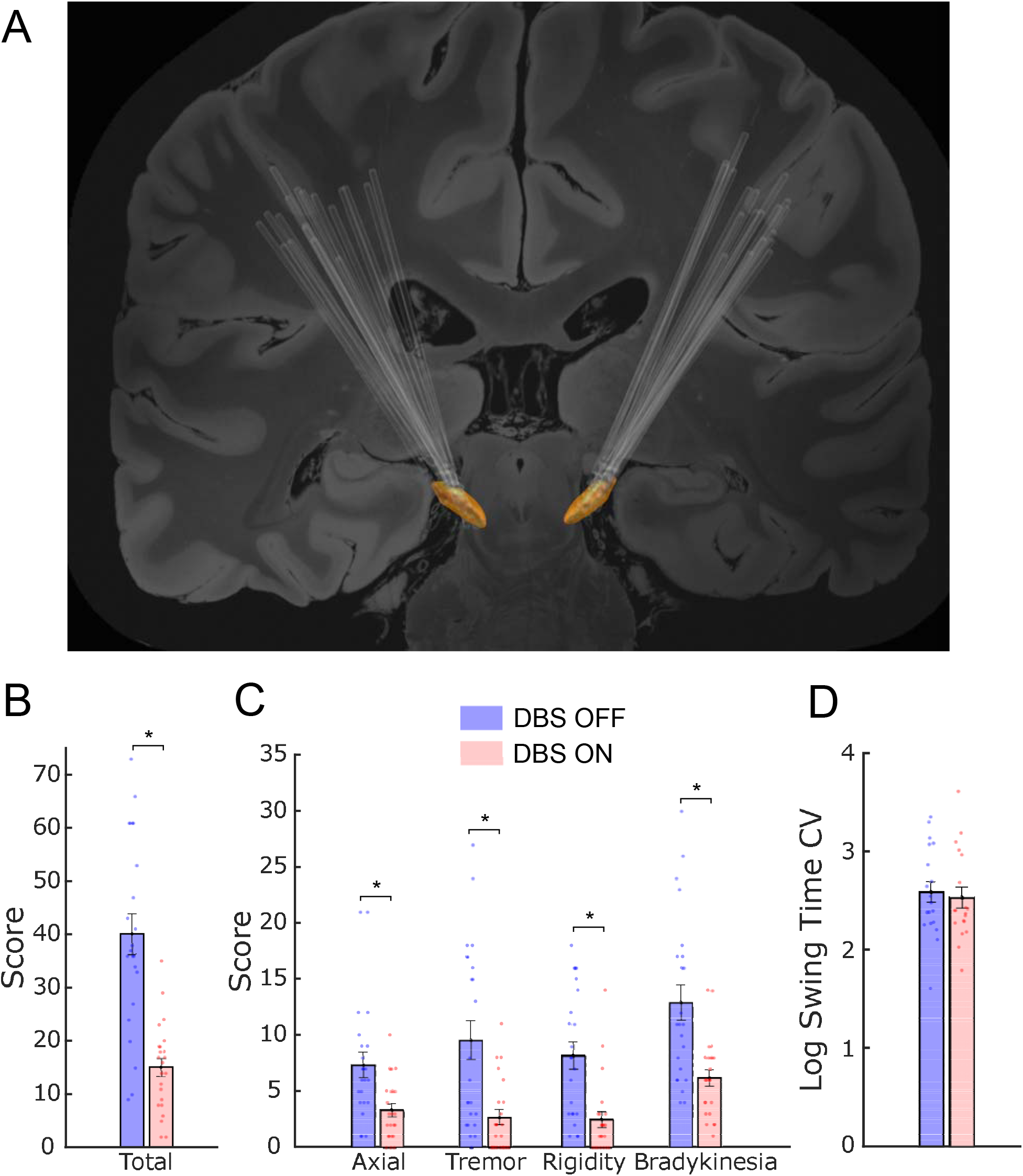
STN DBS improved cardinal motor signs but did not improve swing time variability. (A) DBS electrode lead placement in the STN (orange) for 19 participants. (B) Comparison of total UPDRS III and (C) its sub-scores OFF (blue) and ON (pink) DBS. STN DBS reduced total UPDRS-III and all of its sub-scores. (D) Comparison of swing time variability during the SIP task OFF and ON DBS. STN DBS did not improve swing time variability. Error bars represent SEM. * indicates significant differences.

Cardinal motor signs all improved ON compared to OFF STN DBS, as shown by the reduced total UPDRS III scores, Fig. 2B (t(22) = 9.64, *p* = 2.33e-9) and all of its sub-scores, Fig. 2C, which included axial (t(22) = 5.00, *p* = 5.27e-5), tremor (t(22)=5.46, *p* = 1.75e-5), rigidity (t(22) = 6.38. *p* = 2.01e-6, and bradykinesia (t(22) = 6.61, *p* = 1.21e-6). In contrast, swing time variability did not improve ON compared to OFF DBS, Fig. 2D (t(18) = 0.89, *p* = 0.39).

### NBM atrophy predicted the decline in swing variability

After three years of continuous STN DBS, a subset of eleven individuals with PD were assessed off all therapy (i.e., off DBS and dopaminergic medication) for changes in the UPDRS III and in swing time variability during the SIP task. The remaining individuals from the original cohort did not participate in the three-year follow-up and therefore were not included in the analysis. A portion of a SIP trace from a representative individual at IP and at the three-year timepoint is depicted in Figure 3A and 3B. This individual demonstrated more arrhythmic stepping at the 3-year timepoint compared to their IP visit. The demonstration of gait deterioration in this individual was supported by the group statistic demonstrating that swing time variability during non-freezing stepping increased (worsened), over time, Fig. 3C (t(10) = 4.42; *p* = 0.001).

**Figure 3.**
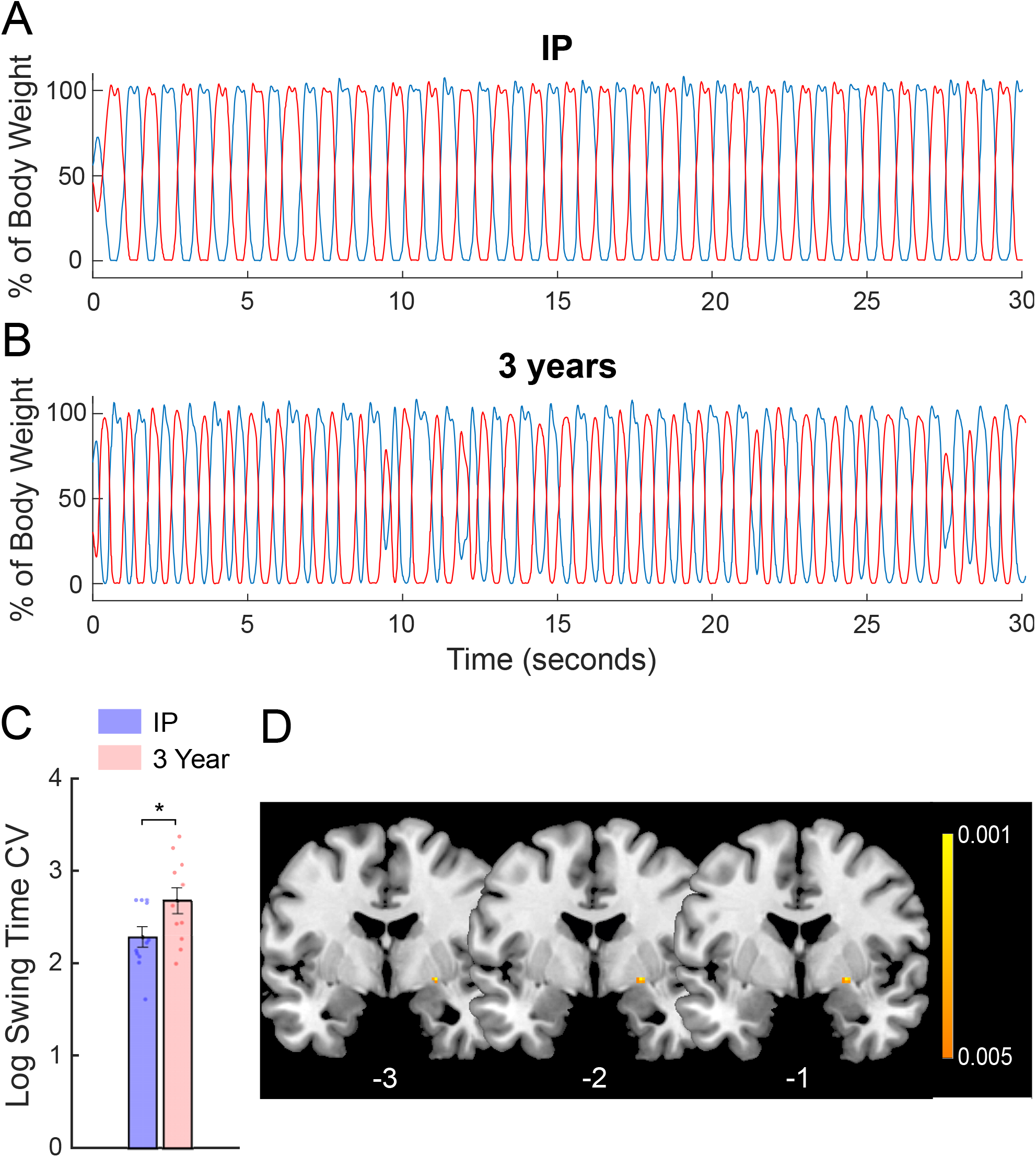
STN DBS improved cardinal motor signs but did not improve swing time variability. (A) DBS electrode lead placement in the STN (orange) for 19 participants. (B) Comparison of total UPDRS III and (C) its sub-scores OFF (blue) and ON (pink) DBS. STN DBS reduced total UPDRS-III and all of its sub-scores. (D) Comparison of swing time variability during the SIP task OFF and ON DBS. STN DBS did not improve swing time variability. Error bars represent SEM. * indicates significant differences.

A multiple regression model found a significant cluster within the NBM where lower gray matter volume was significantly associated with greater increase of swing time variability from IP to three years, Fig. 3D (*p* = 0.001; uncorrected). Information on this cluster is shown in Table 3. Meanwhile, no clusters showed a positive correlation within the NBM between gray matter volume and change in swing time variability over time, nor was there any association between the amygdala and progression of swing time variability.

**Table 3.**
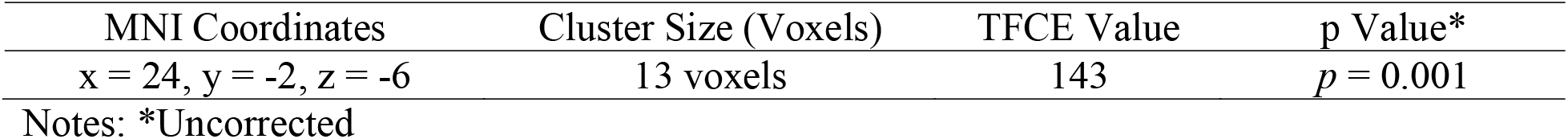
**Correlation between NBM Gray Matter Volume and Worsening of Swing Time Variability at 3 Years**

| MNI Coordinates | Cluster Size (Voxels) | TFCE Value | p Value* |
| --- | --- | --- | --- |
| x = 24, y = -2, z = -6 | 13 voxels | 143 | $p = 0.001$ |

Notes: \*Uncorrected

### Continuous DBS stalled progression of cardinal motor signs measured off therapy

In contrast to the worsening of swing time variability over time, there was no change in UPDRS III, off therapy, between IP and the three-year timepoint, Figure 4A (t(10) = 0.36, *p* = 0.73). This held true for all sub-scores, Fig. 4B, including axial (t(10) = 0.54, *p* = 0.60), tremor (t(10) = 1.31, *p* = 0.22), rigidity (t(10) = 1.27, *p* = 0.23), and bradykinesia (t(10) = 0.89, *p* = 0.40) indicating that there was no observable decline in cardinal motor signs over time, off therapy, after three years of continuous STN DBS.

**Figure 4.**
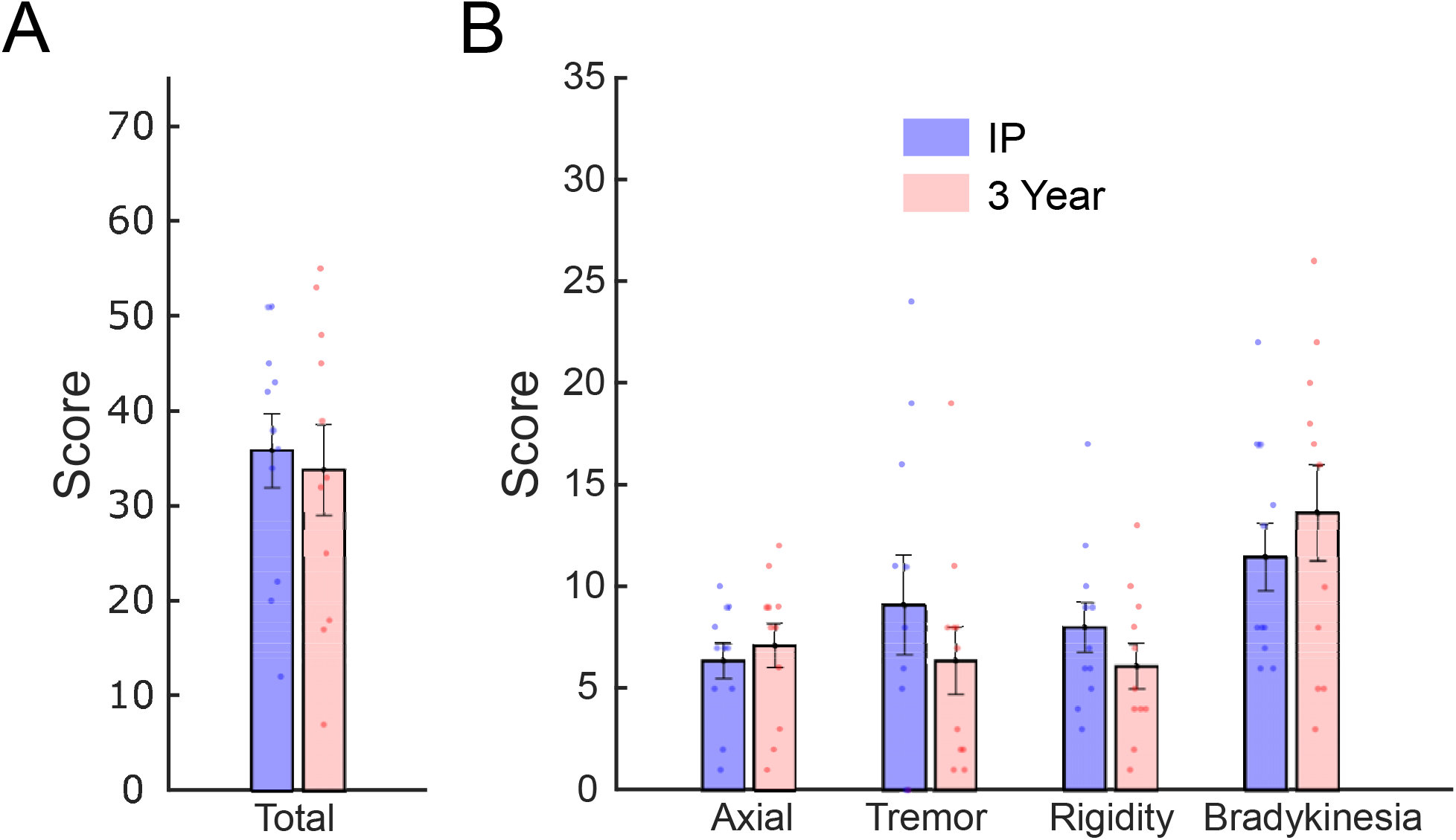
Cardinal motor signs did not worsen over time, measured off therapy, after three years of continuous STN DBS. (A) Total UPDRS III and (B) its sub-scores at IP (blue) and 3 years later (pink) tested off of all therapy. Error bars represent SEM.

## Discussion

The current study investigated the relationship between gray matter volume of the Nucleus Basalis of Meynert (NBM), the central node in the cortical cholinergic system, and gait impairment in individuals with PD through a combination of structural imaging, quantitative gait analysis, and STN DBS. We demonstrated, for the first time, that atrophy of the NBM was associated with greater swing time variability in individuals with PD. STN DBS, which targets the degenerating dopaminergic system in PD, did not improve deficits in swing time variability despite overall improvements in tremor, rigidity, bradykinesia, and axial motor function. It was evident that, off therapy, swing time variability continued to worsen over three years, and the amount of atrophy of NBM predicted the degree of worsening of swing time variability. In contrast, there was no progression of cardinal motor signs, off therapy, after three years of continuous STN DBS. Together, these results point towards a critical role of the cortical cholinergic system in higher order aspects of gait control, such as gait variability in PD, and give insight into the limitations of STN DBS in ameliorating specific domains of gait impairment.

### The influence of the cholinergic system on gait variability

Gait variability is a metric that requires higher-order control and differentiates PD from healthy controls: although both healthy controls and individuals with PD will decrease speed in response to a demanding dual task, only PD show increases in gait variability, such as swing time variability (Yogev et al., 2005). This suggests that individuals with PD show deficits in the automaticity of gait, and increasingly rely on cognitive-related functions to compensate. Additionally, greater gait variability during walking is associated with greater sensorimotor activity, suggesting a role of top-down processing (Berger et al., 2019; Kurz et al., 2012). The stepping-in-place task used here, may be especially effective at highlighting the higher-order contributions to gait variability considering optic flow is minimized, so sensory-related guidance of stepping is more limited compared to forward walking. It has been shown that increased gait variability is associated with impairment in certain cognitive domains in PD, some of which may be related to the cholinergic system (Amboni et al., 2008; Morris et al., 2017; Nantel et al., 2012; Yao et al., 2017). However, there has been no link between gait variability and the cholinergic system until now, despite theoretical predictions (Morris et al., 2019). The results of this study demonstrate for the first time that atrophy of the NBM, the key node in the cortical cholinergic network, is related to greater (i.e., worse) swing time variability in PD, thus providing evidence that the cholinergic system plays a significant role in the underlying mechanism contributing to impairment of higher-order aspects of gait.

The observed relationship between the cholinergic system and swing time variability continues to build on the evidence that PD is not merely a hypodopaminergic disease, but rather a multisystem neurodegenerative disorder (Bohnen et al., 2013). This points to the reliance of higher-order aspects of gait (stepping control) on the cholinergic system, possibly through its effects on attention and other cognitive processes that may become increasingly critical as the automaticity of gait becomes more impaired. Previously, it has been shown that levels of cortical ACh were related to gait speed during forward walking in PD (Bohnen et al., 2013; Rochester et al., 2012). However, gait speed is also intertwined with the dopaminergic network as both levodopa and STN DBS consistently improve gait speed (Ferrarin et al., 2005; Roper et al., 2016; Stolze et al., 2001). We show that swing time variability, a gait variability metric independent of gait speed, is related to the integrity of a key node in the cortical cholinergic network. We propose therefore that a compromised cholinergic system negatively impacts higher-order aspects of gait control. This idea is supported by pharmacological evidence showing that administration of acetylcholinesterase inhibitors improved both attention (Wesnes et al., 2005) and step variability in PD (Henderson et al., 2016). Within this framework, the cholinergic system appears to be the common denominator in the interplay between higher order control of gait and cognition, leaving both processes susceptible to decline in PD as the NBM deteriorates.

Swing time variability is not the only metric of gait variability. Step time, step length, and stride time variability are also all different accepted metrics of gait variability and may also be related to the cholinergic system (Henderson et al., 2016). However, swing time variability is the only speed-independent measure of gait variability of this group (Beauchet et al., 2009; Callisaya et al., 2010; Frenkel-Toledo et al., 2005). We chose to use this as our measure of gait variability in order to limit potential confounds of the effect of DBS or natural aging on gait variability due to changes in speed and better separate from speed-related contributions of the dopaminergic system.

Although the NBM is the key node in the cortical cholinergic system, the pedunculopontine nucleus (PPN) provides cholinergic input at the level of the brainstem and the thalamus and is implicated in gait impairment in PD (Jenkinson et al., 2009; Lau et al., 2015; Thevathasan et al., 2018). Impaired cholinergic integrity of the PPN is strongly related to falls and posture (Müller et al., 2013), and its position in the brainstem allows it to serve as a final outflow block influencing gait (Lewis and Shine, 2016). Whereas the PPN modulates bottom-up processing (Kim et al., 2017), the cortical cholinergic system strongly correlates with cognitive deficits (Bohnen et al., 2015, 2012) and other higher-order top-down processes (Kim et al., 2019). It is also possible that the two systems interact at the level of the thalamus or brainstem to impact aspects of gait and postural control (Hallanger et al., 1987; Parent et al., 1988).

### Differentiating the networks involved in cardinal motor signs and gait variability in PD with STN DBS

STN DBS improved the cardinal motor signs of PD but did not improve swing time variability; swing time variability in individuals with PD was also related to atrophy of the NBM, and was significantly worse than that observed in age-matched controls. These results shed insight into the inconsistency of the efficacy of STN DBS on different metrics of gait variability and swing time asymmetry (Anidi et al., 2018; Blin et al., 1991; Bryant et al., 2011; Curtze et al., 2015; Lord et al., 2011; Rochester et al., 2017). Gait is a highly complex process that encompasses motor and non-motor mechanisms, including executive function, attention, and visuospatial processing (Amboni et al., 2013). STN DBS may successfully address certain motor aspects of gait such as gait speed in PD by targeting the basal ganglia dopaminergic network; however, it would not be expected to address aspects of gait control that rely more heavily on other networks. The lack of improvement in swing time variability from neuromodulation of dopaminergic networks, demonstrates that the control of higher order or non-motor aspects of gait is strongly influenced by other networks, such as the cholinergic network.

### Long-term progression of gait variability and cardinal motor signs

Swing time variability, measured after withdrawal of STN DBS and medication, continued to worsen over three years, whereas cardinal motor signs off therapy did not decline. This result provides further evidence that the worsening of swing time variability over time is a manifestation of progressing neuropathology in non-dopaminergic networks. Other measures of gait variability, such as step time and step length, have also shown worsening over time despite dopaminergic therapy (Rochester et al., 2017). It is unlikely that the observed worsening of swing time variability in the current study is merely due to aging as previous findings in a cohort of older healthy adults showed no worsening of swing time variability, as measured by SD, over a three year period (Rochester et al., 2017), while we observed a 17% worsening in our PD cohort.

Continuous STN DBS appeared to alter the expected decline of cardinal motor signs, measured off all therapy, over three years. This finding builds on previous work from our lab that continuous neuromodulation of the sensorimotor network via STN DBS may alter the progression of cardinal motor signs (Trager et al., 2016). Any disease-modifying effect of neuromodulation of the dopaminergic network would not extend to the cholinergic network and therefore would not modify the progression of higher-order aspects of gait reliant on cholinergic function.

### NBM atrophy predicts worsening of gait variability over time

The degree of atrophy of the NBM predicted the increase in (worsening of) swing time variability three years after initial evaluation. It has been shown that atrophy of the NBM and its microstructure predicted cognitive decline several years later in PD (Pereira et al., 2020; Ray et al., 2018; Schulz et al., 2018). Considering that NBM degeneration occurs early on in the disease process in PD (Braak et al., 2003; Shimada et al., 2009), there is a growing need to find ways of intervening before any accumulating effects start significantly impacting behavior, especially cognition. The link between NBM, cognition, and gait may point towards a potential role for gait variability as a readily accessible metric that will serve as a “canary in the coal mine” to predict impairment in cortical cholinergic networks and subsequent decline in higher-order control of gait, and to serve as a measure of efficacy of novel therapeutic interventions. Such a metric could be crucial to allow for identification of patients who may benefit from early interventions to prevent consequences of increasing gait variability, such as freezing of gait and falls (Morris et al., 2019).

## Limitations

The number of participants in this study, particularly for the IP to 3-year comparison was small. This cohort was limited by the fact that these individuals were part of a larger trial using an investigative sensing neurostimulator (Activa™ PC+S, Medtronic PLC, FDA Investigational Device Exemption (IDE) approved). This is the largest cohort of individuals with this stimulator at any individual site in the world. The cohort examined here also had a relatively long disease duration at the time of their MRI scan (8.1 ± 3.2 years). It is unclear whether individuals with PD earlier in their disease state would demonstrate similar NBM atrophy. However, it is significant that atrophy was already present despite no clear cognitive deficits during the DBS preoperative evaluation. Another possible limitation is that different Parkinsonian signs may return at different rates after discontinuing STN DBS. For instance, axial symptoms return more gradually than tremor (Temperli et al., 2003). During the DBS visit and at the three-year follow-up, DBS was turned off at least 15 minutes prior to testing; we have shown that beta band power, a neural biomarker of hypokinetic aspects of PD, returned to baseline within one minute after DBS was turned off (Trager et al., 2016). Axial symptoms respond much faster when going from the OFF to ON state (Temperli et al., 2003), so it is unlikely that the ON DBS condition represented a sub-optimal stimulation state.

The SIP task provides a safe environment for assessment of gait due to the presence of a harness, has been validated as a measure of gait impairment in PD, and minimizes the role of visual feedback. One drawback is that one cannot measure speed or step length, which have both been associated with the cholinergic system, since there is no forward motion component to the task. However, the goal of this study was to measure a marker of gait variability independent of speed to better separate from possible dopaminergic contributions. Another limitation is that the analyses were restricted to a region of interest analysis rather than a whole-brain analysis due to the small size of the NBM. However, this was an a priori decision based on previous findings for the NBM’s role in cognitive decline in PD. We found no associations between swing time variability and our control region of interest, the amygdala. Finally, there was no follow-up MRI to track the change in NBM atrophy alongside the decline in swing time variability or MRIs for the age-matched control cohort. Future studies should combine both quantitative metrics of gait with longitudinal MRIs to further investigate the predictive power of the NBM for gait impairment and vice versa.

## Conclusions

This study demonstrated that deficits in swing time variability, a measure of gait variability that is independent of gait speed, in PD were related to a degenerating cortical cholinergic system: greater swing time variability was associated with lower gray matter volume in the NBM, the key node in the cortical cholinergic network. Swing time variability was greater in the PD cohort compared to a group of age-matched controls and was not associated with atrophy of the amygdala, a similar sized region in the brain. Neuromodulation of the dopaminergic basal ganglia-cortical network, via STN DBS, improved cardinal motor signs of tremor, rigidity, bradykinesia, and axial motor function, but did not improve swing time variability. Furthermore, swing time variability continued to decline over three years of continuous STN DBS, and pre-operative NBM atrophy was a predictor of worsening swing time variability. In contrast, after three years of continuous STN DBS, cardinal motor signs measured off therapy were no different from those prior to activation of the DBS system. The above results support a framework in which higher-order aspects of gait control may be reliant on cortical cholinergic processing. Swing time variability is a readily accessible measure of impairment that could provide insight into the health of cortical cholinergic networks in PD and could identify individuals, who may benefit from early interventions to prevent the devastating behavioral manifestations of the degeneration of the cortical cholinergic system over the course of PD.

## Data Availability

Data will be made available on request when possible.

## Acknowledgements

We would like to thank Dr. Jaimie Henderson, the members of the Human Motor Control and Neuromodulation lab, Varsha Prabhakar and Talora Martin for their help in collecting the control data, and, most importantly, the participants who dedicated their time to this study.

## Authors’ Roles

KBW: Conceptualization; Methodology; Software; Validation; Formal Analysis; Writing – Original Draft; Visualization; Funding Acquisition

JEP: Conceptualization; Data Curation; Writing - Review & Editing; Project administration

HMBS: Conceptualization; Validation; Resources; Writing – Review & Editing; Supervision; Project Administration; Funding acquisition

## Data Access and Responsibility

HMBS takes responsibility for the integrity of the data and the accuracy of the data analysis. Data will be made available on request when possible.

## Abbreviations

ACh: Acetylcholine
CV: Coefficient of Variation
DBS: Deep brain stimulation
FWE: Family-wise error
IP: Initial Programming
NBM: Nucleus Basalis of Meynert
PD: Parkinson’s Disease
PPN: Pedunculopontine nucleus
SIP: Stepping-In-Place
STN: Subthalamic Nucleus
UPDRS: Unified Parkinson’s Disease Rating Scale

## References

Amboni, M., Barone, P., Hausdorff, J.M., 2013. Cognitive contributions to gait and falls: Evidence and implications. Mov. Disord. https://doi.org/10.1002/mds.25674

Amboni, M., Cozzolino, A., Longo, K., Picillo, M., Barone, P., 2008. Freezing of gait and executive functions in patients with Parkinson’s disease. Mov. Disord. https://doi.org/10.1002/mds.21850

Anidi, C., O’Day, J.J., Anderson, R.W., Afzal, M.F., Syrkin-Nikolau, J., Velisar, A., Bronte-Stewart, H.M., 2018. Neuromodulation targets pathological not physiological beta bursts during gait in Parkinson’s disease. Neurobiol. Dis. 120, 107–117. https://doi.org/10.1016/j.nbd.2018.09.004

Arcolin, I., Corna, S., Giardini, M., Giordano, A., Nardone, A., Godi, M., 2019. Proposal of a new conceptual gait model for patients with Parkinson’s disease based on factor analysis. Biomed. Eng. Online. https://doi.org/10.1186/s12938-019-0689-3

Avants, B.B., Epstein, C.L., Grossman, M., Gee, J.C., 2008. Symmetric diffeomorphic image registration with cross-correlation: Evaluating automated labeling of elderly and neurodegenerative brain. Med. Image Anal. https://doi.org/10.1016/j.media.2007.06.004

Ballinger, E.C., Ananth, M., Talmage, D.A., Role, L.W., 2016. Basal Forebrain Cholinergic Circuits and Signaling in Cognition and Cognitive Decline. Neuron. https://doi.org/10.1016/j.neuron.2016.09.006

Barrett, M.J., Sperling, S.A., Blair, J.C., Freeman, C.S., Flanigan, J.L., Smolkin, M.E., Manning, C.A., Druzgal, T.J., 2019. Lower volume, more impairment: Reduced cholinergic basal forebrain grey matter density is associated with impaired cognition in Parkinson disease. J. Neurol. Neurosurg. Psychiatry. https://doi.org/10.1136/jnnp-2019-320450

Beauchet, O., Annweiler, C., Lecordroch, Y., Allali, G., Dubost, V., Herrmann, F.R., Kressig, R.W., 2009. Walking speed-related changes in stride time variability: Effects of decreased speed. J. Neuroeng. Rehabil. https://doi.org/10.1186/1743-0003-6-32

Berger, A., Horst, F., Steinberg, F., Thomas, F., Müller-Eising, C., Schöllhorn, W.I., Doppelmayr, M., 2019. Increased gait variability during robot-assisted walking is accompanied by increased sensorimotor brain activity in healthy people. J. Neuroeng. Rehabil. https://doi.org/10.1186/s12984-019-0636-3

Blin, O., Ferrandez, A.M., Pailhous, J., Serratrice, G., 1991. Dopa-sensitive and Dopa-resistant gait parameters in Parkinson’s disease. J. Neurol. Sci. https://doi.org/10.1016/0022-510X(91)90283-D

Bohnen, N.I., Albin, R.L., Müller, M.L.T.M., Petrou, M., Kotagal, V., Koeppe, R.A., Scott, P.J.H., Frey, K.A., 2015. Frequency of cholinergic and caudate nucleus dopaminergic deficits across the predemented cognitive spectrum of parkinson disease and evidence of interaction effects. JAMA Neurol. https://doi.org/10.1001/jamaneurol.2014.2757

Bohnen, N.I., Frey, K.A., Studenski, S., Kotagal, V., Koeppe, R.A., Scott, P.J.H., Albin, R.L., Müller, M.L.T.M., 2013. Gait speed in Parkinson disease correlates with cholinergic degeneration. Neurology. https://doi.org/10.1212/WNL.0b013e3182a9f558

Bohnen, N.I., Kaufer, D.I., Hendrickson, R., Ivanco, L.S., Lopresti, B.J., Constantine, G.M., Mathis, C.A., Davis, J.G., Moore, R.Y., DeKosky, S.T., 2006. Cognitive correlates of cortical cholinergic denervation in Parkinson’s disease and parkinsonian dementia. J. Neurol. https://doi.org/10.1007/s00415-005-0971-0

Bohnen, N.I., Müller, M.L.T.M., Kotagal, V., Koeppe, R.A., Kilbourn, M.R., Gilman, S., Albin, R.L., Frey, K.A., 2012. Heterogeneity of cholinergic denervation in Parkinson’s disease without dementia. J. Cereb. Blood Flow Metab. https://doi.org/10.1038/jcbfm.2012.60

Braak, H., Del Tredici, K., Rüb, U., De Vos, R.A.I., Jansen Steur, E.N.H., Braak, E., 2003. Staging of brain pathology related to sporadic Parkinson’s disease. Neurobiol. Aging. https://doi.org/10.1016/S0197-4580(02)00065-9

Brontë-Stewart, H., Louie, S., Batya, S., Henderson, J.M., 2010. Clinical motor outcome of bilateral subthalamic nucleus deep-brain stimulation for Parkinson’s disease using image-guided frameless stereotaxy. Neurosurgery. https://doi.org/10.1227/NEU.0b013e3181ecc887

Bryant, M.S., Rintala, D.H., Hou, J.G., Charness, A.L., Fernandez, A.L., Collins, R.L., Baker, J., Lai, E.C., Protas, E.J., 2011. Gait variability in Parkinson’s disease: Influence of walking speed and dopaminergic treatment. Neurol. Res. https://doi.org/10.1179/1743132811Y.0000000044

Callisaya, M.L., Blizzard, L., Schmidt, M.D., McGinley, J.L., Srikanth, V.K., 2010. Ageing and gait variability-a population-based study of older people. Age Ageing. https://doi.org/10.1093/ageing/afp250

Curtze, C., Nutt, J.G., Carlson-Kuhta, P., Mancini, M., Horak, F.B., 2015. Levodopa Is a Double-Edged Sword for Balance and Gait in People With Parkinson’s Disease. Mov. Disord. https://doi.org/10.1002/mds.26269

Ewert, S., Plettig, P., Li, N., Chakravarty, M.M., Collins, D.L., Herrington, T.M., Kühn, A.A., Horn, A., 2018. Toward defining deep brain stimulation targets in MNI space: A subcortical atlas based on multimodal MRI, histology and structural connectivity. Neuroimage. https://doi.org/10.1016/j.neuroimage.2017.05.015

Ferrarin, M., Rizzone, M., Bergamasco, B., Lanotte, M., Recalcati, M., Pedotti, A., Lopiano, L., 2005. Effects of bilateral subthalamic stimulation on gait kinematics and kinetics in Parkinson’s disease. Exp. Brain Res. https://doi.org/10.1007/s00221-004-2036-5

Frenkel-Toledo, S., Giladi, N., Peretz, C., Herman, T., Gruendlinger, L., Hausdorff, J.M., 2005. Effect of gait speed on gait rhythmicity in Parkinson’s disease: Variability of stride time and swing time respond differently. J. Neuroeng. Rehabil. https://doi.org/10.1186/1743-0003-2-23

Gang, M., Baba, T., Hosokai, Y., Nishio, Y., Kikuchi, A., Hirayama, K., Hasegawa, T., Aoki, M., Takeda, A., Mori, E., Suzuki, K., 2020. Clinical and Cerebral Metabolic Changes in Parkinson’s Disease With Basal Forebrain Atrophy. Mov. Disord. https://doi.org/10.1002/mds.27988

Goetz, C.G., Fahn, S., Martinez-Martin, P., Poewe, W., Sampaio, C., Stebbins, G.T., Stern, M.B., Tilley, B.C., Dodel, R., Dubois, B., Holloway, R., Jankovic, J., Kulisevsky, J., Lang, A.E., Lees, A., Leurgans, S., LeWitt, P.A., Nyenhuis, D., Olanow, C.W., Rascol, O., Schrag, A., Teresi, J.A., Van Hilten, J.J., LaPelle, N., 2007. Movement disorder society-sponsored revision of the unified Parkinson’s disease rating scale (MDS-UPDRS): Process, format, and clinimetric testing plan. Mov. Disord. https://doi.org/10.1002/mds.21198

Gratton, C., Yousef, S., Aarts, E., Wallace, D.L., D’Esposito, M., Silver, M.A., 2017. Cholinergic, But Not Dopaminergic or Noradrenergic, Enhancement Sharpens Visual Spatial Perception in Humans. J. Neurosci. https://doi.org/10.1523/JNEUROSCI.2405-16.2017

Hallanger, A.E., Levey, A.I., Lee, H.J., Rye, D.B., Wainer, B.H., 1987. The origins of cholinergic and other subcortical afferents to the thalamus in the rat. J. Comp. Neurol. https://doi.org/10.1002/cne.902620109

Henderson, E.J., Lord, S.R., Brodie, M.A., Gaunt, D.M., Lawrence, A.D., Close, J.C.T., Whone, A.L., Ben-Shlomo, Y., 2016. Rivastigmine for gait stability in patients with Parkinson’s disease (ReSPonD): a randomised, double-blind, placebo-controlled, phase 2 trial. Lancet Neurol. https://doi.org/10.1016/S1474-4422(15)00389-0

Horn, A., Kühn, A.A., 2015. Lead-DBS: A toolbox for deep brain stimulation electrode localizations and visualizations. Neuroimage. https://doi.org/10.1016/j.neuroimage.2014.12.002

Horn, A., Li, N., Dembek, T.A., Kappel, A., Boulay, C., Ewert, S., Tietze, A., Husch, A., Perera, T., Neumann, W.J., Reisert, M., Si, H., Oostenveld, R., Rorden, C., Yeh, F.C., Fang, Q., Herrington, T.M., Vorwerk, J., Kühn, A.A., 2019. Lead-DBS v2: Towards a comprehensive pipeline for deep brain stimulation imaging. Neuroimage. https://doi.org/10.1016/j.neuroimage.2018.08.068

Husch, A.V. Petersen, M., Gemmar, P., Goncalves, J., Hertel, F., 2018. PaCER - A fully automated method for electrode trajectory and contact reconstruction in deep brain stimulation. NeuroImage Clin. https://doi.org/10.1016/j.nicl.2017.10.004

Jenkinson, N., Nandi, D., Muthusamy, K., Ray, N.J., Gregory, R., Stein, J.F., Aziz, T.Z., 2009. Anatomy, physiology, and pathophysiology of the pedunculopontine nucleus. Mov. Disord. https://doi.org/10.1002/mds.22189

Johnston, M. V., McKinney, M., Coyle, J.T., 1979. Evidence for a cholinergic projection to neocortex from neurons in basal forebrain. Proc. Natl. Acad. Sci. U. S. A. https://doi.org/10.1073/pnas.76.10.5392

Kim, K., Müller, M.L.T.M., Bohnen, N.I., Sarter, M., Lustig, C., 2019. The cortical cholinergic system contributes to the top-down control of distraction: Evidence from patients with Parkinson’s disease. Neuroimage. https://doi.org/10.1016/j.neuroimage.2017.12.012

Kim, K., Müller, M.L.T.M., Bohnen, N.I., Sarter, M., Lustig, C., 2017. Thalamic cholinergic innervation makes a specific bottom-up contribution to signal detection: Evidence from Parkinson’s disease patients with defined cholinergic losses. Neuroimage. https://doi.org/10.1016/j.neuroimage.2017.02.006

Klinkenberg, I., Sambeth, A., Blokland, A., 2011. Acetylcholine and attention. Behav. Brain Res. https://doi.org/10.1016/j.bbr.2010.11.033

Kurz, M.J., Wilson, T.W., Arpin, D.J., 2012. Stride-time variability and sensorimotor cortical activation during walking. Neuroimage. https://doi.org/10.1016/j.neuroimage.2011.08.084

Lau, B., Welter, M.L., Belaid, H., Fernandez Vidal, S., Bardinet, E., Grabli, D., Karachi, C., 2015. The integrative role of the pedunculopontine nucleus in human gait. Brain. https://doi.org/10.1093/brain/awv047

Lewis, S.J.G., Shine, J.M., 2016. The Next Step: A Common Neural Mechanism for Freezing of Gait. Neuroscientist. https://doi.org/10.1177/1073858414559101

Lord, S., Baker, K., Nieuwboer, A., Burn, D., Rochester, L., 2011. Gait variability in Parkinson’s disease: An indicator of non-dopaminergic contributors to gait dysfunction? J. Neurol. https://doi.org/10.1007/s00415-010-5789-8

Mesulam, M. □Marsel, Mufson, E.J., Levey, A.I., Wainer, B.H., 1983. Cholinergic innervation of cortex by the basal forebrain: Cytochemistry and cortical connections of the septal area, diagonal band nuclei, nucleus basalis (Substantia innominata), and hypothalamus in the rhesus monkey. J. Comp. Neurol. https://doi.org/10.1002/cne.902140206

Mesulam, M.M., 2013. Cholinergic circuitry of the human nucleus basalis and its fate in Alzheimer’s disease. J. Comp. Neurol. https://doi.org/10.1002/cne.23415

Morris, R., Lord, S., Lawson, R.A., Coleman, S., Galna, B., Duncan, G.W., Khoo, T.K., Yarnall, A.J., Burn, D.J., Rochester, L., 2017. Gait Rather Than Cognition Predicts Decline in Specific Cognitive Domains in Early Parkinson’s Disease. Journals Gerontol. - Ser. A Biol. Sci. Med. Sci. https://doi.org/10.1093/gerona/glx071

Morris, R., Martini, D.N., Madhyastha, T., Kelly, V.E., Grabowski, T.J., Nutt, J., Horak, F., 2019. Overview of the cholinergic contribution to gait, balance and falls in Parkinson’s disease. Park. Relat. Disord. https://doi.org/10.1016/j.parkreldis.2019.02.017

Mufson, E.J., Martin, T.L., Mash, D.C., Wainer, B.H., Mesulam, M.M., 1986. Cholinergic projections from the parabigeminal nucleus (Ch8) to the superior colliculus in the mouse: a combined analysis of horseradish peroxidase transport and choline acetyltransferase immunohistochemistry. Brain Res. https://doi.org/10.1016/0006-8993(86)91114-5

Müller, M.L.T.M., Albin, R.L., Kotagal, V., Koeppe, R.A., Scott, P.J.H., Frey, K.A., Bohnen, N.I., 2013. Thalamic cholinergic innervation and postural sensory integration function in Parkinson’s disease. Brain. https://doi.org/10.1093/brain/awt247

Nakano, I., Hirano, A., 1984. Parkinson’s disease: Neuron loss in the nucleus basalis without concomitant Alzheimer’s disease. Ann. Neurol. https://doi.org/10.1002/ana.410150503

Nantel, J., de Solages, C., Bronte-Stewart, H., 2011. Repetitive stepping in place identifies and measures freezing episodes in subjects with Parkinson’s disease. Gait Posture. https://doi.org/10.1016/j.gaitpost.2011.05.020

Nantel, J., McDonald, J.C., Tan, S., Bronte-Stewart, H., 2012. Deficits in visuospatial processing contribute to quantitative measures of freezing of gait in Parkinson’s disease. Neuroscience. https://doi.org/10.1016/j.neuroscience.2012.07.007

Panisset, M., 2004. Freezing of gait in Parkinson’s disease. Neurol. Clin. https://doi.org/10.1016/j.ncl.2004.05.004

Parent, A., Paré, D., Smith, Y., Steriade, M., 1988. Basal forebrain cholinergic and noncholinergic projections to the thalamus and brainstem in cats and monkeys. J. Comp. Neurol. https://doi.org/10.1002/cne.902770209

Pereira, J.B., Hall, S., Jalakas, M., Grothe, M.J., Strandberg, O., Stomrud, E., Westman, E., van Westen, D., Hansson, O., 2020. Longitudinal degeneration of the basal forebrain predicts subsequent dementia in Parkinson’s disease. Neurobiol. Dis. https://doi.org/10.1016/j.nbd.2020.104831

Ray, N.J., Bradburn, S., Murgatroyd, C., Toseeb, U., Mir, P., Kountouriotis, G.K., Teipel, S.J., Grothe, M.J., 2018. In vivo cholinergic basal forebrain atrophy predicts cognitive decline in de novo Parkinson’s disease. Brain. https://doi.org/10.1093/brain/awx310

Rochester, L., Galna, B., Lord, S., Yarnall, A.J., Morris, R., Duncan, G., Khoo, T.K., Mollenhauer, B., Burn, D.J., 2017. Decrease in Aβ42 predicts dopa-resistant gait progression in early Parkinson disease. Neurology. https://doi.org/10.1212/WNL.0000000000003840

Rochester, L., Yarnall, A.J., Baker, M.R., David, R. V., Lord, S., Galna, B., Burn, D.J., 2012. Cholinergic dysfunction contributes to gait disturbance in early Parkinson’s disease. Brain. https://doi.org/10.1093/brain/aws207

Rogers, J.D., Brogan, D., Mirra, S.S., 1985. The nucleus basalis of Meynert in neurological disease: A quantitative morphological study. Ann. Neurol. https://doi.org/10.1002/ana.410170210

Roper, J.A., Kang, N., Ben, J., Cauraugh, J.H., Okun, M.S., Hass, C.J., 2016. Deep brain stimulation improves gait velocity in Parkinson’s disease: a systematic review and meta-analysis. J. Neurol. https://doi.org/10.1007/s00415-016-8129-9

Schulz, J., Pagano, G., Fernández Bonfante, J.A., Wilson, H., Politis, M., 2018. Nucleus basalis of Meynert degeneration precedes and predicts cognitive impairment in Parkinson’s disease. Brain. https://doi.org/10.1093/brain/awy072

Shimada, H., Hirano, S., Shinotoh, H., Aotsuka, A., Sato, K., Tanaka, N., Ota, T., Asahina, M., Fukushi, K., Kuwabara, S., Hattori, T., Suhara, T., Irie, T., 2009. Mapping of brain acetylcholinesterase alterations in Lewy body disease by PET. Neurology. https://doi.org/10.1212/WNL.0b013e3181ab2b58

Smith, S.M., Nichols, T.E., 2009. Threshold-free cluster enhancement: Addressing problems of smoothing, threshold dependence and localisation in cluster inference. Neuroimage. https://doi.org/10.1016/j.neuroimage.2008.03.061

Smulders, K., Dale, M.L., Carlson-Kuhta, P., Nutt, J.G., Horak, F.B., 2016. Pharmacological treatment in Parkinson’s disease: Effects on gait. Park. Relat. Disord. https://doi.org/10.1016/j.parkreldis.2016.07.006

Stolze, H., Klebe, S., Poepping, M., Lorenz, D., Herzog, J., Hamel, W., Schrader, B., Raethjen, J., Wenzelburger, R., Mehdorn, H.M., Deuschl, G., Krack, P., 2001. Effects of bilateral subthalamic nucleus stimulation on parkinsonian gait. Neurology. https://doi.org/10.1212/WNL.57.1.144

Temperli, P., Ghika, J., Villemure, J.G., Burkhard, P.R., Bogousslavsky, J., Vingerhoets, F.J.G., 2003. How do parkinsonian signs return after discontinuation of subthalamic DBS? Neurology. https://doi.org/10.1212/WNL.60.1.78

Thevathasan, W., Debu, B., Aziz, T., Bloem, B.R., Blahak, C., Butson, C., Czernecki, V., Foltynie, T., Fraix, V., Grabli, D., Joint, C., Lozano, A.M., Okun, M.S., Ostrem, J., Pavese, N., Schrader, C., Tai, C.H., Krauss, J.K., Moro, E., 2018. Pedunculopontine nucleus deep brain stimulation in Parkinson’s disease: A clinical review. Mov. Disord. https://doi.org/10.1002/mds.27098

Trager, M.H., Koop, M.M., Velisar, A., Blumenfeld, Z., Nikolau, J.S., Quinn, E.J., Martin, T., Bronte-Stewart, H., 2016. Subthalamic beta oscillations are attenuated after withdrawal of chronic high frequency neurostimulation in Parkinson’s disease. Neurobiol. Dis. https://doi.org/10.1016/j.nbd.2016.08.003

Wesnes, K.A., McKeith, I., Edgar, C., Emre, M., Lane, R., 2005. Benefits of rivastigmine on attention in dementia associated with Parkinson disease. Neurology. https://doi.org/10.1212/01.wnl.0000184517.69816.e9

Yao, Z., Shao, Y., Han, X., 2017. Freezing of gait is associated with cognitive impairment in patients with Parkinson disease. Neurosci. Lett. https://doi.org/10.1016/j.neulet.2017.07.004

Yogev, G., Giladi, N., Peretz, C., Springer, S., Simon, E.S., Hausdorff, J.M., 2005. Dual tasking, gait rhythmicity, and Parkinson’s disease: Which aspects of gait are attention demanding? Eur. J. Neurosci. https://doi.org/10.1111/j.1460-9568.2005.04298.x

Yttri, E.A., Dudman, J.T., 2018. A Proposed Circuit Computation in Basal Ganglia: History-Dependent Gain. Mov. Disord. https://doi.org/10.1002/mds.27321

Zaborszky, L., Hoemke, L., Mohlberg, H., Schleicher, A., Amunts, K., Zilles, K., 2008. Stereotaxic probabilistic maps of the magnocellular cell groups in human basal forebrain. Neuroimage. https://doi.org/10.1016/j.neuroimage.2008.05.055

